# Deep learning for polygenic prediction: The role of heritability, interaction type and sample size

**DOI:** 10.1101/2024.10.25.24316156

**Authors:** Jason Grealey, Gad Abraham, Guillaume Méric, Rodrigo Cánovas, Martin Kelemen, Shu Mei Teo, Agus Salim, Michael Inouye, Yu Xu

## Abstract

Polygenic scores (PGS), which aggregate the effects of genetic variants to estimate predisposition for a disease or trait, have potential clinical utility in disease prevention and precision medicine. Recently, there has been increasing interest in using deep learning (DL) methods to develop PGS, due to their strength in modelling complex non-linear relationships (such as GxG) that conventional PGS methods may not capture. However, the perceived value of DL for polygenic scores is unclear. In this study, we assess the underlying factors impacting DL performance and how they can be better utilised for PGS development. We simulate large-scale realistic genotype-to-phenotype data, with varying genetic architectures of phenotypes under quantitative control of three key components: (a) total heritability, (b) variant-variant interaction type, and (c) proportion of non-additive heritability. We compare the performance of one of most common DL methods (multi-layer perceptron, MLP) on varying training sample sizes, with two well-established PGS methods: a purely additive model (pruning and thresholding, P+T) and a machine learning method (Elastic net, EN). Our analyses show EN has consistently better overall performance across traits of different architectures and training data of different sizes. However, MLP saw the largest performance improvements as sample size increases. MLP outperformed P+T for most traits and achieves comparable performance as EN for numerous traits at the largest sample size assessed (N=100k), suggesting DL may offer some advantages in future when they can be trained on biobanks of millions of samples. We further found that one-hot encoding of variant input can improve performance of every method, particularly for traits with non-additive variance. Overall, we show how different underlying factors impact how well methods leverage non-additivity for polygenic prediction.

## Introduction

Polygenic scores (PGS), which aggregate the effects of many genetic variants into a single number, have become an important tool to predict the genetic predisposition of an individual towards a phenotype and have been shown to have promising utility such as in disease prevention and precision medicine^1–3^. There is increasing interest in using deep learning (DL) approaches to develop PGS of complex traits^4–11^. Known as universal function approximators^12,13^, the value of deep learning models is in their ability to model complex non-linear effects among genetic variants and their flexibility in combining with other non-genetic factors for subsequent applications (e.g. disease related biomarkers and environmental factors for disease risk models).

Human traits, including quantitative traits and diseases, are heritable to varying degrees and many of them have been found to have a highly polygenic architecture (i.e., their variance is accounted for by many thousands or even millions of genetic variants genome-wide)^14^. While studies have shown that for most phenotypes^15,16^ the associated variants contribute largely in a linearly additive manner, non-linear interaction effects (GxG) are present and sometimes make a substantial contribution to the genetic variation of phenotypes, e.g. autoimmune diseases^7,17^.

It has been shown that common machine learning methods, such as elastic net and gradient boosting trees, can capture GxG in the genetic prediction of common traits and diseases^5,18,19^, frequently improving PGS performance. While these methods do not explicitly model interaction terms, GxG can still be captured to an extent through variant encoding or inherently non-linear structures of the model. Deep learning methods readily model complex non-linear relationships and have recently been proposed for PGS development of various human traits^4– 7,20,21^. DL methods have been found to improve PGS of several traits and diseases, such as breast cancer^6^, Alzheimer’s disease^10^ and type 1 diabetes^7^, but in many cases substantially improved performance over simpler machine learning models has not been found^4,5,7^. DL methods may also be susceptible to confounding by joint tagging effects, whereby GxG is in fact attributed to unaccounted additive genetic variants, and only provide moderate improvements in prediction performance even under extreme genetic architectures.^22^

Despite substantial efforts, it remains unclear under what conditions (if at all) DL may offer an advantage over simple approaches to polygenic score construction. Here, we investigate how and to what extent key factors of genetic architecture and sample size affect the performance of PGS models and in particular, under what circumstances DL methods outperform linear models. To answer these questions, we simulated genotype data of 100,000 individuals with realistic linkage disequilibrium (LD) patterns, and phenotypes whose genetic architectures were of varying: (a) total heritability (broad sense), (b) types of GxG interaction, and (c) proportion of non-additive heritability. We compared the performance of a suite of common methods for PGS development of these simulated phenotypes, which included a univariate linear model (pruning and thresholding), a regularized linear regression (elastic net), and a deep learning approach (multi-layer perceptron). We also investigated the impact of training data sizes and variant encoding types on the performance of these methods. Our findings inform study designs and methodology selection for future PGS development.

## Methods

### Simulating genotypes

HAPGEN2^23^ was used to simulate genotypes with realistic linkage disequilibrium (LD) patterns. As an empirical reference panel from which to draw haplotypes, we utilised chromosome 22 (171,457 variants in total) from 99 individuals of the phase 3 of the 1000 Genomes project^24^ (Finnish subset). This reference panel was used to generate 100,000 simulated individual haplotypes which after conversion to genotypes contained 100,455 variants (after keeping variants with minor allele frequency (MAF) between 1% and 40%). No variant was found to violate the Hardy-Weinberg equilibrium (*p*<10^−6^) using PLINK^25^.

### Simulating phenotypes

Phenotypes (in this study, continuous traits) were simulated using the simulated genotypes above, where genotypes were coded in a minor allele dosage format {0, 1, 2}. For each phenotype, a total of 1,000 variants were randomly chosen (∼1% of the total variants in the simulated dataset) and were given an effect size randomly drawn from a normal distribution with a standard deviation σ_β_ (others have effect sizes of 0):

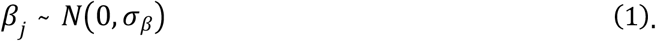

After all the 1,000 variants were given a linear effect size, as drawn from equation (1) with σ_β_ initialised at 0.01, these effect sizes were used to scale the non-additive heritability for the trait. Of these 1,000 causal variants, if the trait was influenced by GxG, 500 of them (250 variant-variant pairs are randomly sampled) were given non-additive effects, which were modelled according to the two locus interaction types from Li and Reich^26^. A non-additive effect was simulated under a given combination of effect alleles for both variants according to four interaction types: threshold (“T”), recessive/recessive (“RR”), exclusive/or (“XOR”), and heterozygote/heterozygote (“HH”, previously named as “m16”^26^) (**Figure 4**). If an individual contains this specific combination of effect alleles, these variants will exhibit a variant interaction effect on their phenotype.

The GxG interaction effect for a given pair of variants *k* of sample *i* is determined by the following equation:

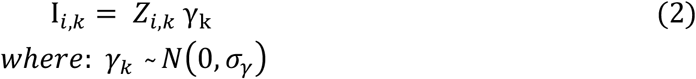

where *Z*_*i,k*_ is an indicator function for the GxG interaction types (**Figure 4**) for interacting variant pair *k* of sample *i* and is either 1 or 0 depending on the combination of genotypes and the GxG interaction type; if *Z*_*i,k*_ = 1 (i.e. a given interaction type exhibits), the interaction effect size is drawn from a normal distribution. σ_*γ*_ is initialised at 0.01 and scaled with respect to the total (i.e. additive and non-additive) genetic variation within the phenotype to control the level of non-additive variation contributing towards the phenotype (See below).

As well as GxG interaction effects and linear effects, there was also a proportion of noise in the phenotype that genetics do not explain, i.e. a non-heritable contribution. This was modelled as follows:

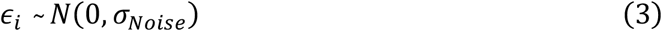

where σ_*Noise*_ is scaled to fix the heritability of the trait.

The above equations combine for the phenotype like so:

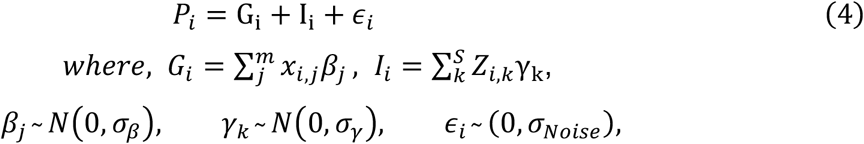

*G*_*i*_, *I*_*i*_ and *ϵ*_*i*_ are the combined linear effects, combined non-additive effects for variants exhibiting interactions and the noise for sample *i* respectively; *x*_*i,j*_ is the number of effect alleles present in variant *j* of sample *i*; *β*_*j*_ is the linear effect size drawn from equation (1) for the effect allele in variant *j*; *Z*_*i,k*_ determines if the *k*^*th*^ pair of interacting variants exhibits a certain GxG interaction type in sample *i* where 4 interaction types are applied using the two locus penetrance tables in this study (**Figure 4**). The phenotype value for sample *i* is determined by summing all contributions from the linear effects of *m* simulated variants, the GxG interaction effects of *S* pairs of simulated interaction variants, and the noise.

### Heritability simulation

After the first step of initialization of phenotype values (i.e. its noise, non-additive and linear components) as described above, we then performed linear regression to scale the contribution of each component to control the non-additive contribution to the total heritability and its total heritability or broad sense heritability for the purpose of simulating phenotypes of different settings^27^.

As described above, the sum of all genetic effects on a given phenotype for individual *i* is as follows:

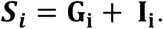

To determine the proportion of non-additive heritability in the total genetic effects, we performed the following linear regression across all individuals:

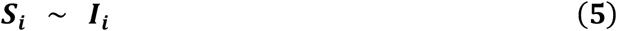

where the goodness-of-fit *γR*^2^) of the regression determines the total non-additive contribution to the heritability. For example, if the *R*^2^ was 20%, then only 80% of the total heritability would be narrow sense (linear additive) and the other 20% being non-additive, i.e. from GxG. These non-additive effects are increased or decreased by scaling all the pairwise interacting effects to obtain the required level of non-additive variance in the trait. The linear regression was performed using Scikit-learn python package^28^.

Similarly, we performed the following regression to determine the broad sense heritability of a given trait:

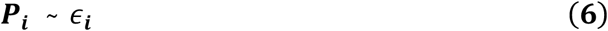

with which, noise in the trait is increased or decreased to obtain the required broad sense heritability.

### Summary of the simulated dataset

In total, we simulated 60 phenotypes of different settings, which were under control of three parameters: (a) total heritability (20, 50, or 80%), (b) GxG interaction type (HH, RR, XOR, or T)^26^, and (c) proportion of non-additive heritability (0, 20, 40, 60, or 80%).

For each phenotype, 500 variants were randomly selected and given a linear contribution for the phenotype. The remaining 500 variants were given linear and paired GxG interaction effects for a given epistatic model, where variants are randomly selected to generate 250 non-overlapping pairs. In the case of no non-additive effects, these 500 variants were only given linear effects. These effects were summed for all 1000 variants, noise was added, and heritability and the proportion of non-additive heritability were fixed. Three predictive models: (i) additive PGS, (ii) elastic net PGS and (iii) feed forward neural networks, were used to develop polygenic scores for these phenotypes, which are detailed below.

Three sample sets of different sizes (100k, 50k, and 10k) were randomly selected from the simulated dataset for each phenotype, each of which was then split 60/40% into training and testing sets (**Figure 1**). Then every prediction model used the same generated sample sets to train PGS models and test their performance.

**Figure 1.**
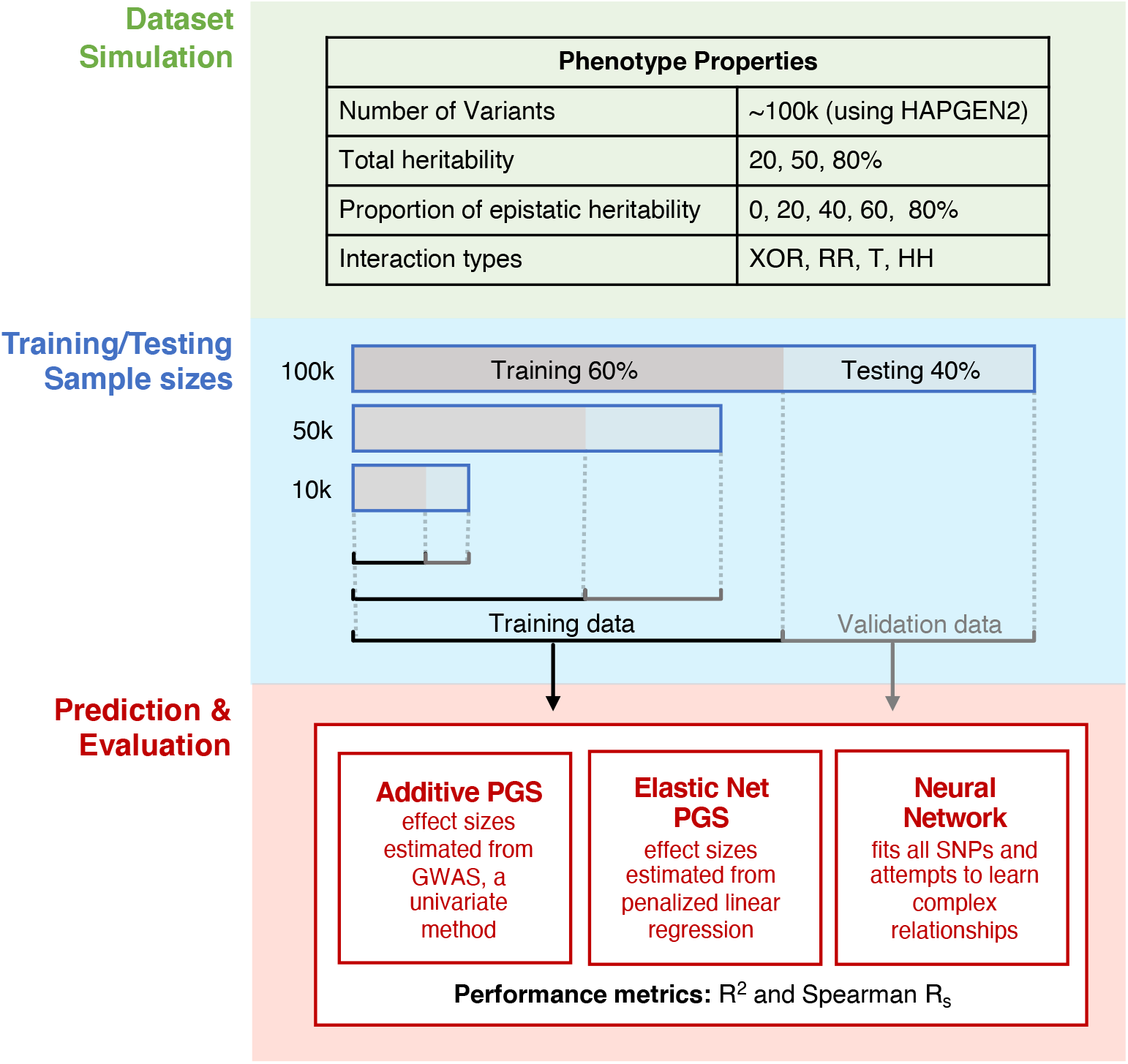
Schematic study design for data simulation and genetic prediction. Simulations of 100k genotypes were generated using HAPGEN2 and subsampled to smaller datasets of 50k and 10k samples. Using these simulated genotypes, traits with different settings of heritability, GxG interaction type, and proportion of non-additive heritability were generated. The samples were split into training and testing sets in each dataset of 100k, 50k and 10k samples, after which they were used to train and test the prediction methods (neural networks, elastic net PGS and additive PGS).

### Additive PGS method P+T

This additive PGS method assumes that the genetic variants have linear additive effects on PGS of the trait, and develops PGS of a trait using the weighted sum of genotypes of the selected variants for that trait:

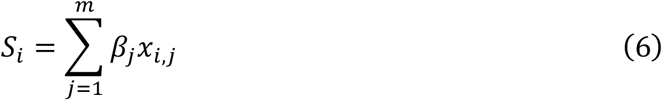

where *S*_i_ is a polygenic score for individual *i*; *x*_*i,j*_ is the genotype dosage of variant *j* of the individual *i*; the *β* _*j*_ is the effect size of the variant *j* that is usually obtained through the univariate statistical association tests on training data; the *m* variants were often selected through a LD pruning/clumping and p-value thresholding step^29^ (so this method is often named as *P+T*). The software PLINK^25^ was used to estimate the univariate effect sizes for each simulated variant on the training data of a given simulated phenotype. Using these univariate estimations, PRSice-2^30^ was then employed to develop PGS of the phenotype on the training data. PRSice-2 performs LD clumping to reduce the correlation amongst variants and then tests thousands of optimised p-value filtered PGSs to obtain the most predictive PGS.

### Elastic net PGS method

Elastic net (EN) also assumes variants have a linear additive effect, estimated via penalised regression, where all the variants are jointly fit together. SparSNP^31^ is a tool designed to fit penalised linear models for genetic prediction, and was used to perform elastic-net regression in this study. SparSNP minimizes the following loss function for estimating the effect sizes of genetic variants:

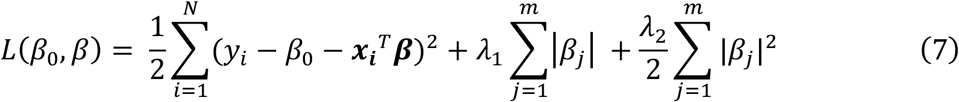

where, *y*_*i*_ is the simulated phenotype value; *x*_*i*_ are the genotype dosages of the *m* variants for sample *i* (i.e. the 100,455 simulated variants after quality control); *β* is the vector of effect sizes for the m variants; *β*_0_ is the intercept term; *λ*_1_ and *λ*_2_ are the penalties for a LASSO and Ridge regularisation respectively. A 5-fold cross validation was performed for 10 times in SparSNP to select the optimal *λ*_1_, *λ*_2_ pair on a given training set, where *λ*_2_was set as 0.2 and *λ*_1_ was identified from a default set of thirty options in SparSNP. Finally, effect sizes of variants were estimated by minimizing equation (7) on the training set, which are then used to construct the PGS using equation (6).

### Neural networks

Multilayered perceptrons (MLPs; also called feed-forward neural networks) are one of most common neural network architectures and can improve genetic prediction of quantitative traits, e.g. blood cell traits^5^. MLPs do not make any assumptions about the distributions behind the data they fit, and can be trained to approximate any smooth function in theory^13^. They usually consist of nodes (functions) connected to many other layers through directed acyclic graphs^13^; the output of a layer is used as the input to subsequent layers and element-wise transformed by non-linear activation functions, which allows for it to model complex correlations. Given *m* nodes in the *Lth* layer, the output of a node *i* in the next or *γL +* 1)*th* layer is calculated like so:

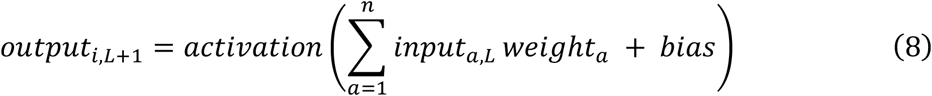

where *output*_*i,L*+1_ is the output of a node *i* in the *γL +*1)*th* layer; *input*_*a,L*_ is the input from node *a* (from the total *n* nodes) in the previous layer. Each *input*_*a,L*_ is multiplied by a weight (i.e. *weight*_*a*_) and added to a *bias*, which are then summed up and passed into an activation function as the output of node *i*. As mentioned above these activation functions are used to incorporate non-linearity to the modelling process. This process occurs from the first hidden layer, to any hidden layers until the output is reached (**Supplementary Figures 13-14**). MLP models were implemented with Keras (keras.io) and TensorFlow^32^ in our study.

### Genotype Encoding

We considered two different types of genotype encoding, additive encoding (i.e. effect allele dosage) and one hot encoding, as the input of the prediction models in this study (**Figure 2**). The first encoding involved the use of Plink v1.9^25^ to encode the variants into allelic dosages, where the variants were input as counts of the effect alleles (i.e. 0, 1, 2). In “one hot” encoding, variants were encoded into the absence or presence of their genotype classes.

**Figure 2.**
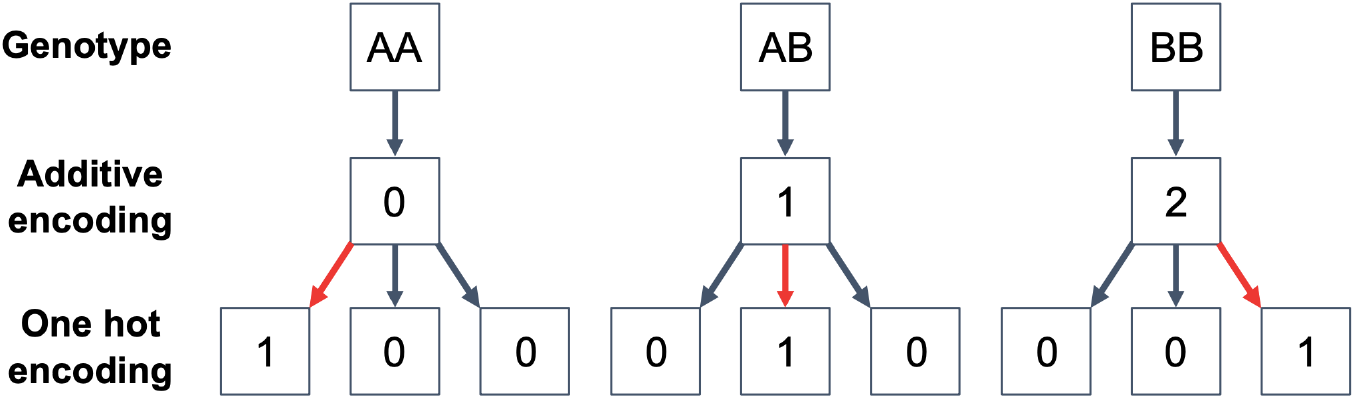
Schematics of genotype encoding. This schematic shows the two different variants encodings used in this study and shows how genotypes are represented in additive and one hot encodings.

### Hyperparameter optimisation

An essential component in neural network model training, in particular MLP in this study, is hyperparameter optimisation. Hyperparameters are variables that dictate the network’s structure, its complexities and training process, which are set before the model training. Each set of hyperparameters can perform differently on a given task, thus a search must be conducted to determine the optimal set for each task. The hyperparameter search was conducted using Talos^33^ package which aids in performing the random search for the best set of hyperparameters for a given task (i.e. predicting each phenotype) on the training data. Given a list of hyperparameters to optimise from, Talos randomly searches this list to create numerous unique combinations of hyperparameters (see details in **Supplementary Table 1**), which were used to determine the best performing set of hyperparameters for a given phenotype.

### Assessment of prediction accuracy

Finally, the two metrics: coefficient of determination (*R*^2^) and Spearman correlation coefficient (Rs), were used to measure the performance of each PGS method on the testing data of any given phenotype setting as described above.

## Results

In this study, we simulated genotype data of 100,000 individuals using the 1000G reference panel, with which we further simulated 60 phenotypes of different settings, including (a) total heritability (20, 50, or 80%), (b) GxG interaction type (HH, RR, XOR, or T)^26^, and (c) proportion of non-additive heritability (0, 20, 40, 60, or 80%). We then evaluated the performance of three polygenic score methods, including a simple additive PGS method (the pruning and thresholding), a linear machine learning method (elastic net) and a deep learning method (multilayered perceptron), in predicting these simulated phenotypes using training data of different sizes. Two different types of genotype encoding: additive dosage encoding and one hot encoding, were also applied to investigate its impact on the performance of PGS methods.

### Performance of polygenic prediction methods across different settings

Across various simulation settings, elastic net performed consistently well across phenotypes and training datasets when compared to the other methods (**Figures 3-4 and Supplementary Figures 1-5**). Using the 10k simulated data (training sample size: 6k), EN improved R^2^ by 22% on average over the simple additive method (P+T) across traits of different GxG interaction types and neural network model MLP underperformed EN by 65% in terms of R^2^ (additive variant encoding for P+T and EN; **Figure 3**). However, at this smallest sample size applied, P+T slightly performed better than EN for a few traits with low heritability and high proportions of non-additive heritability, e.g. P+T improves over EN by 3% (R^2^) for traits of 20% heritability and 60% non-additive heritability (additive variant encoding; **Supplementary Figure 1** and **Supplementary Table 2**). As the sample size increased, EN continued to outperform both P+T and MLP for most traits. EN is commonly used with additive variant encoding (‘dosage model’), and the MLP with one-hot encoding frequently outperformed this approach. In particular, for traits with HH interactions, MLP had R^2^ 25% greater than EN with additive variant encoding when sample size was 100k. However, using one hot encoding enabled EN to outperform MLP (**Figure 3a**).

**Figure 3.**
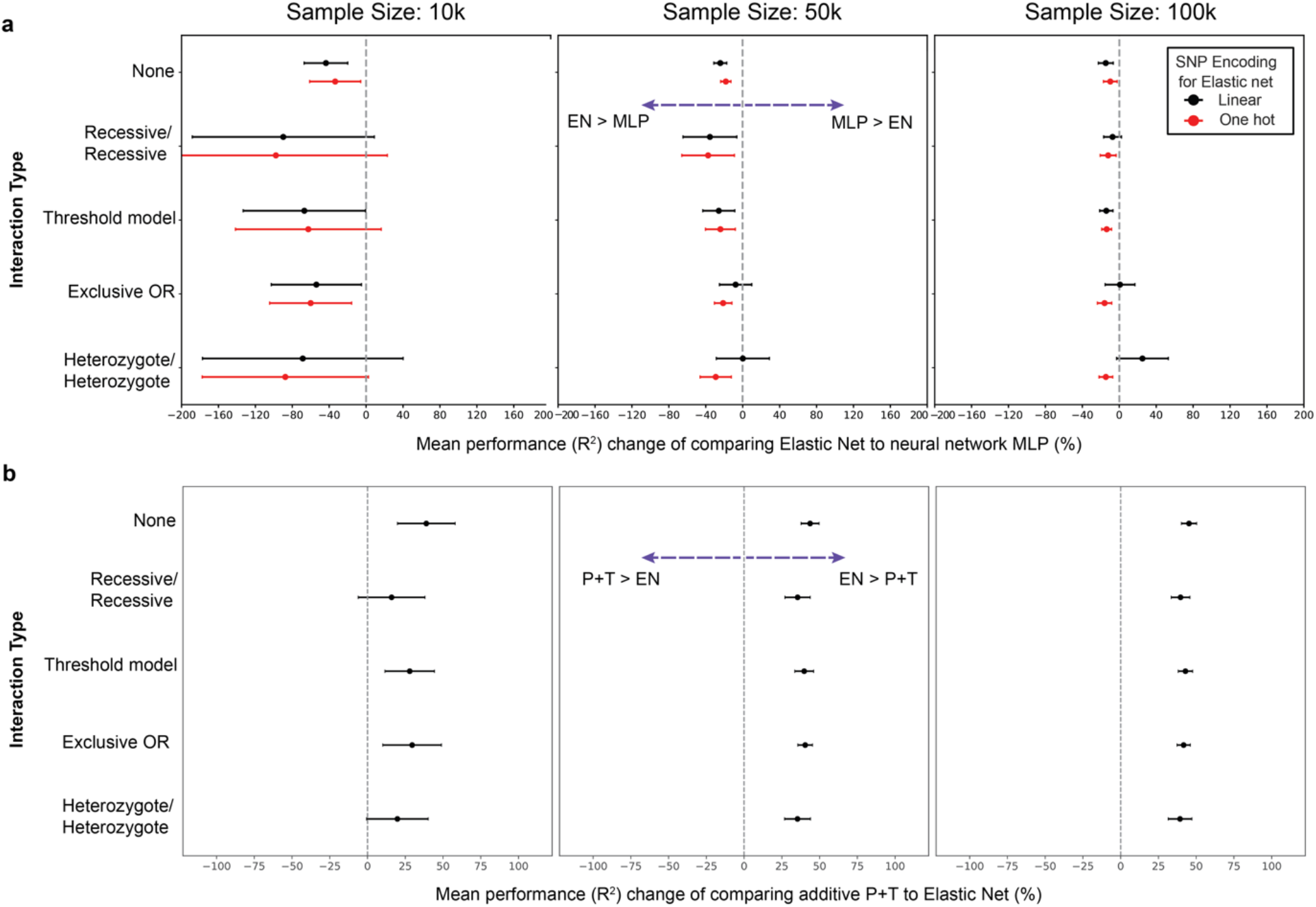
Performance comparison of different PGS methods. a. Elastic net is more accurate than neural network MLP in PGS development. Each sub-plot shows the percentage change in the predictive performance (R^2^) between Elastic net and MLP at a given sample size. Simulated traits are grouped by interaction and variant encoding types, then we compare the performance between Elastic net and MLP by using the mean and standard deviation of 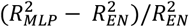 in a selected trait group, where 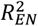 is the R^2^ performance of EN on a trait. Note that both the one hot (red) and additively (black) encoded elastic net PGS methods are compared against one hot encoded MLP. **b. Elastic net outperforms additive P+T method**. Each sub-plot shows the percentage change in the predictive performance (R^2^) of additive PGS method P+T and elastic net at a given total sample size, which are measured using the mean and standard deviation of 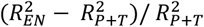 in each trait group by GxG interaction type.

**Figure 4.**
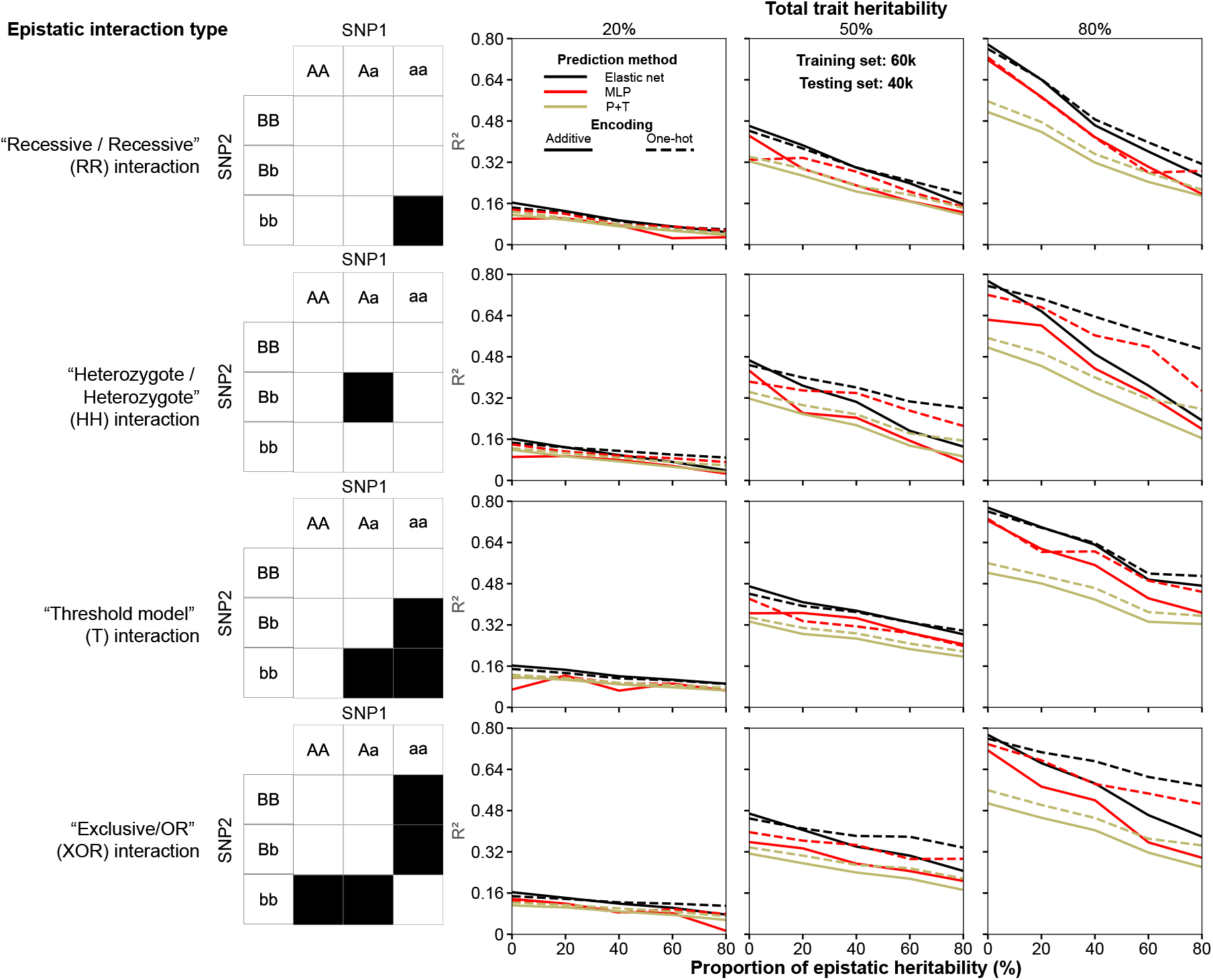
Heritability and proportion of non-additive heritability affect prediction performances of different PGS methods (sample size: 100k). Predictive performance (R^2^) of all the three PGS methods with the two variant encoding types were compared for traits of different groups, where elastic net PGS are in black, neural network MLP in red and additive P+T in yellow; solid lines and dashed lines represent additively and one hot encoded models respectively. The plots detail each PGS method’s performance for a given phenotype. Each row has the same underlying interaction model labelled by the tables and each column has the same total heritability as noted by the column title and within each sub-plot, the *x*-axis details the proportion of non-linear contribution for a given trait.

### One hot encoding of variants frequently improved polygenic prediction

We further assessed how using one-hot encoding as variant inputs affect the accuracy of the three PGS methods. Overall, relative to additive encoding (‘dosage model’), we found one-hot encoding improved the predictive accuracy (R^2^) for traits with non-additive variance on average by 42% for MLP, 14% for EN, and 20% for additive P+T PGS method (**Figures 3a, 4-5** and **Supplementary Figures 6-7**). However, for purely additive traits, one-hot encoding resulted in a percentage change in R^2^ performance of +14%, -9% and +8%, when using MLP, EN and P+T respectively. This indicates that one hot encoding will consistently increase MLP performance but may worsen EN’s performance, depending on the genetic architecture (which is rarely known *a priori*). Note that (i) the MLP using additive encoding were only tested on data with total sample size of 100k due to its poor performance on smaller data sets and extremely expensive training costs (both significant time and computational resource needed), and (ii) performance gains at relatively low heritability (20%) can be susceptible to substantial noise and should be interpreted with caution.

**Figure 5.**
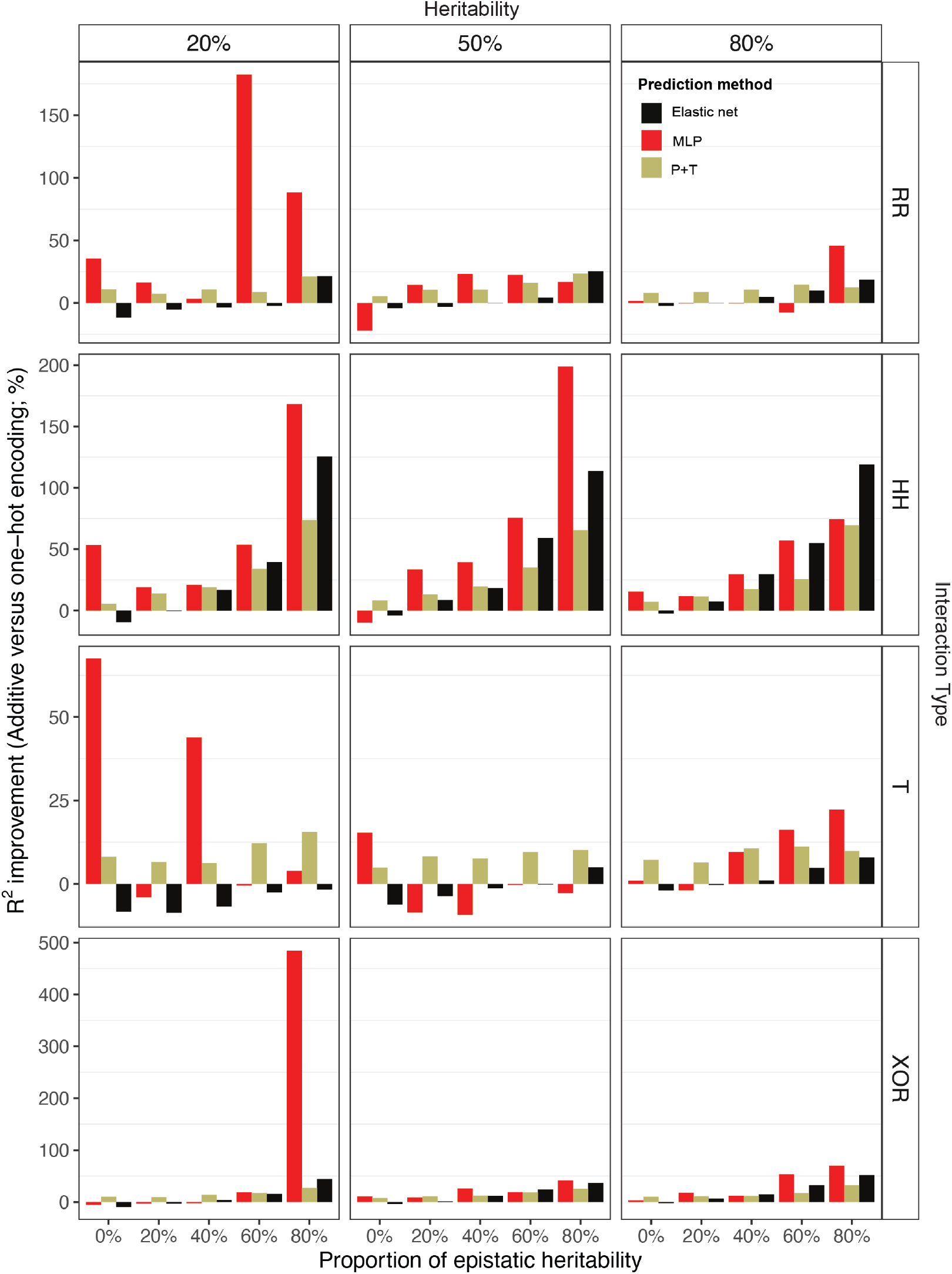
R^2^ performance improvement of PGS methods using the one-hot encoding over the additive encoding for traits of different groups (sample size: 100k). Each row has the same interaction model as noted by the row title and each column has the same total heritability as noted by the column title and within each sub-plot, the *x*-axis shows the proportion of non-linear contribution for a given trait. The improvement is calculated using 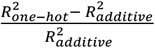, where 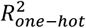 is the R^2^ performance of a PGS method using one-hot encoding for a given trait.

**Figure 6.**
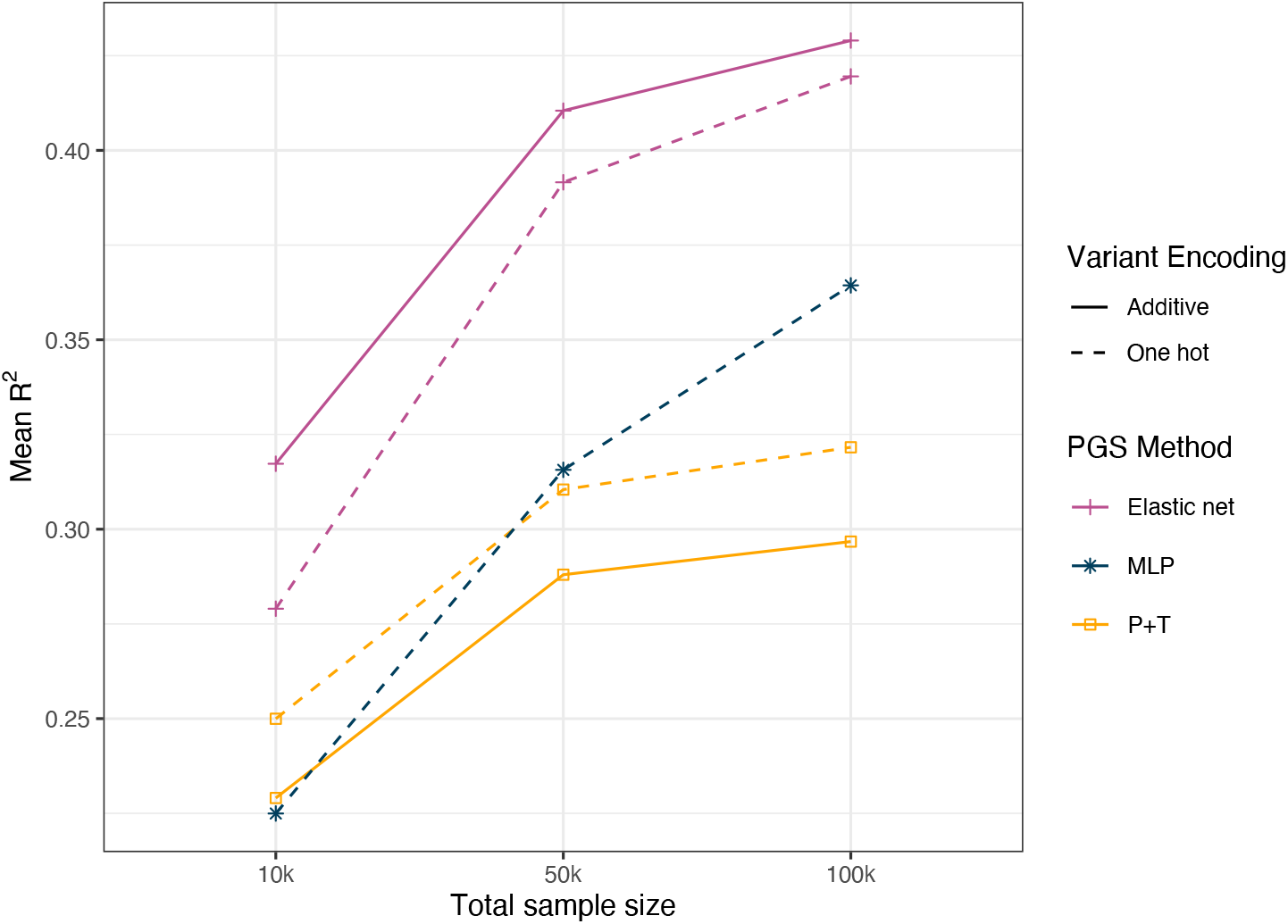
The R^2^ performance improvements of PGS methods by sample size increase. This plot shows the mean R^2^ performance of each PGS method (P+T, elastic net and MLP) with either of the two variant encoding types across these simulated traits with 50% total heritability and either no or 20% non-linear contribution at given sample sizes (10k, 50k, 100k).

### Sample size and relative performance of polygenic prediction method

We examined how increasing the sample size of training data results in an increased predictive power for each PGS method (**Figure 6, Supplementary Figure 8**). One hot encoded MLP saw the largest increase in predictive accuracy: increasing the sample size from 10k to 100k yielded a 62% increase in mean R^2^ scores for traits with 50% heritability and 20% or less GxG (0.22 to 0.36), compared to less than 50% for other PGS methods (**Figure 6**). Across all 60 simulated traits, the MLP’s mean R^2^ improved by 113% (0.15 to 0.32), while other methods showed less than 56% improvement (**Supplementary Figure 8)**. Similarly, increasing the sample size from 50k to 100k resulted in the MLP achieving 15% mean R^2^ improvement for traits with 50% heritability and 20% or less GxG, and a 19% improvement across all simulated traits. In contrast, other methods showed less than a 7% increase for both trait groups. In addition, one hot encoding allowed EN to gain larger improvements when sample size increases compared with using additive variant encoding. For example, increasing from 10k to 50k, the one hot encoded EN had a mean R^2^ increase of 42% (from 0.24 to 0.34), but the additively encoded EN saw a smaller improvement of 29% across all simulated traits.

### Elastic net PGS compared to P+T

Consistent with previous studies, our results showed that elastic net outperformed the additive P+T method for almost every trait under different settings (**Figure 4, Supplementary Figures 1-5**). For example, EN consistently outperformed P+T for all the traits (mean R^2^ improvement: 40%) when a larger sample size (50k and 100k) was used in training; even with the smallest sample size (10k), there were only 10 traits (out of 60) that had a lower R^2^ score with EN.

We further examined how differently EN and P+T methods estimate linear effect sizes of variants in PGS development for traits of different settings. Overall, our results showed the outperformance of EN can be reflected in its better estimation of linear effect sizes of causal variants of the trait (both additively encoded and sample size = 100k; **Supplementary Figures 9-12**). For example, for purely additive traits, the Spearman correlation (R_s_) between the true linear effect size (from simulation) and the EN-estimated effect size was about 25% higher on average when compared with using P+T method (**Supplementary Figure 10**). When total heritability of the trait decreases, EN maintained its accuracy in estimating variant effect sizes (R_s_ = 0.71, 0.73 and 0.72 for traits with total heritability of 80%, 50% and 20% respectively), but P+T saw a significant performance drop (R_s_ = 0.63, 0.56 and 0.54 for traits with total heritability of 80%, 50% and 20% respectively) (**Supplementary Figure 10**). We also found EN was able to better capture true linear effect sizes of variants for traits that are controlled by very high or low proportions of GxG interactions **(Supplementary Figures 9, 11-12**). For example, the R_s_ between the true linear effect size and the estimated effect size from EN was 43% higher on average for traits with either 20% or 80% of non-linear heritability (80% total heritability; **Supplementary Figures 9, 11**) than additive P+T, but this improvement decreased to 9% for traits with 50% of non-linear heritability (**Supplementary Figure 12)**.

## Discussion

In this work, we simulated realistic genotype-to-phenotype data with varying key parameters then compared the performance of deep learning using a multi-layer perceptron to two other common methods (i.e. elastic net and P+T, pruning and thresholding). These key parameters included GxG interaction types, total trait heritability, proportions of non-linear heritability (i.e. from GxG), genotype encoding (additive and one hot encoding), and different sample sizes for training data. Our results showed that traits with low GxG heritability were best predicted by EN but when the proportion of GxG increases, one hot encoding allowed EN to outperform MLP. Our results also showed that the MLP performed considerably better with an increased sample size in training, and as the total trait heritability increases, the relative performance of MLP in comparison with the linear PGS methods increased. Our results suggest that as the size of the training dataset increases substantially beyond 100k toward a million or more individuals, neural network models may achieve equal or better performance as linear PGS methods. However, currently the computational and financial expense of training even an MLP to UK Biobank data is out of reach for the vast majority of academic groups. As costs come down, neural network models, such as MLP, could be useful for the prediction of highly heritable, substantially GxG phenotypes (e.g. some autoimmune diseases) in massive-scale biobanks, e.g. those of millions of individuals.

We found that EN was better at capturing the true linear effect sizes present in the causal variants involved in GxG, indicating EN can better predict traits with GxG even when no interaction terms are explicitly defined in the model. When individual-level genotypes are not available, lasso and related models can be run on GWAS summary statistics using tools such as LDpred^34,35^, Lassosum^36^, PRS-CS^37^ and SBayesRC^38^. Studies have also shown some traits may benefit from PGS methods (e.g. EN) that are based on individual-level genetic data in the current era of large-scale biobanks such as blood cell traits^39^, and ensemble PGS methods, that combine both summary-level (or PGS previously developed in external cohorts) and individual-level data, can result in improved PGS for phenotypes such as coronary heart disease^40^. Nonetheless, our findings support the use of and continued access to individual-level data by *bona fide* researchers so that optimal PGS can be constructed using the most advanced methods.

## Limitations

Whilst the MLP model used in our work is relatively standard and unspecialized, domain knowledge, such as total heritability and GxG interaction type, can be utilised to further optimize neural network architectures. These factors may make neural networks become better predictive models in polygenic prediction of certain traits. Our study utilized simulated phenotypes involving statistical GxG, with a fixed proportion of variance attributed to epistasis. However, in real data analyses, the presence of statistical epistasis can be confounded by LD. In such cases, untyped causal variants may be jointly tagged by SNPs included in the dataset, potentially manifesting as statistical epistasis^41^. To differentiate between these joint tagging effects and true epistasis, dedicated methods are required.^22^ In this study, we only included GxG interaction types that were enumerated by one of the previous studies^26^, a variation of diverse pairwise or two-loci interactions. However, it is possible that higher orders of interactions, not considered in this study, could be present within the human genome. For instance, higher order of interactions have been reported in genes affecting several non-human traits, such as chicken body weight^42^ or colony morphology in yeast^43^. If such interactions exist in humans as well, it is conceivable that neural networks or other complex prediction methods would be more favourable in their polygenic prediction. Finally, we could not justify the costs (both financial and carbon emissions) of simulating data and training neural networks to datasets substantially greater than 100k individuals. We believe such an approach may be justifiable for real data for select autoimmune diseases where substantial GxG is likely (e.g. type 1 diabetes); however, given the paucity of autoimmune cases in existing biobanks a concerted effort would be needed to assemble and harmonise individual-level data for neural network training.

## Conclusion

In summary, this work provides a detailed assessment of neural network models for predicting traits in diverse genetic architectures, in comparison with two commonly used linear PGS methods. It gives general insights into the application of deep learning methods in polygenic prediction, and provides guidance for the selection of optimal PGS methods, variant encoding approach, and training sample size when developing PGS for a target trait. Investigations into customised neural network models, that utilise the genetic architecture of a target trait, may represent a promising future for deep learning in polygenic prediction.

## Data Availability

All data produced in the present study are available upon reasonable request to the authors

## Carbon impact of this study

Based in Victoria, Australia, the computational methods used in this study had an estimated carbon footprint of 2,973 kgCO_2_, which is equivalent to 3,130 tree months. This was estimated using calculated using green-algorithms.org v1.0^44^.

## Code availability

The original codes used to simulate phenotypes of various genetic architectures are available at https://github.com/JasonGrealey/Simulations. The codes of using the three methods (P+T, EN and MLP) to develop PGS are available at https://github.com/xuyu-cam/Deep-learning-for-genetic-prediction-of-complex-traits.

## Conflicts of Interest

M.I. is a trustee of the Public Health Genomics (PHG) Foundation, a member of the Scientific Advisory Board of Open Targets, and has research collaborations with AstraZeneca, Nightingale Health and Pfizer which are unrelated to this study.

## Acknowledgements

This study was supported by the Victorian Government’s Operational Infrastructure Support (OIS) program. JG was supported by a La Trobe University Postgraduate Research Scholarship jointly funded by the Baker Heart and Diabetes Institute and a La Trobe University Full-Fee Research Scholarship.

The support of the UK Economic and Social Research Council (ESRC) is gratefully acknowledged (ES/T013192/1). This work was supported by core funding from: the UK Medical Research Council (MR/L003120/1), the British Heart Foundation (RG/13/13/30194; RG/18/13/33946) and the NIHR Cambridge Biomedical Research Centre (BRC-1215-20014). This work was also supported by Health Data Research UK, which is funded by the UK Medical Research Council, Engineering and Physical Sciences Research Council, Economic and Social Research Council, Department of Health and Social Care (England), Chief Scientist Office of the Scottish Government Health and Social Care Directorates, Health and Social Care Research and Development Division (Welsh Government), Public Health Agency (Northern Ireland), British Heart Foundation, and Wellcome. This study was supported by the Victorian Government’s Operational Infrastructure Support (OIS) program. The views expressed are those of the authors and not necessarily those of the NHS, the NIHR or the Department of Health and Social Care. M.I. was supported by the Munz Chair of Cardiovascular Prediction and Prevention.

## Supplementary Figures

**Supplementary figure 1.**
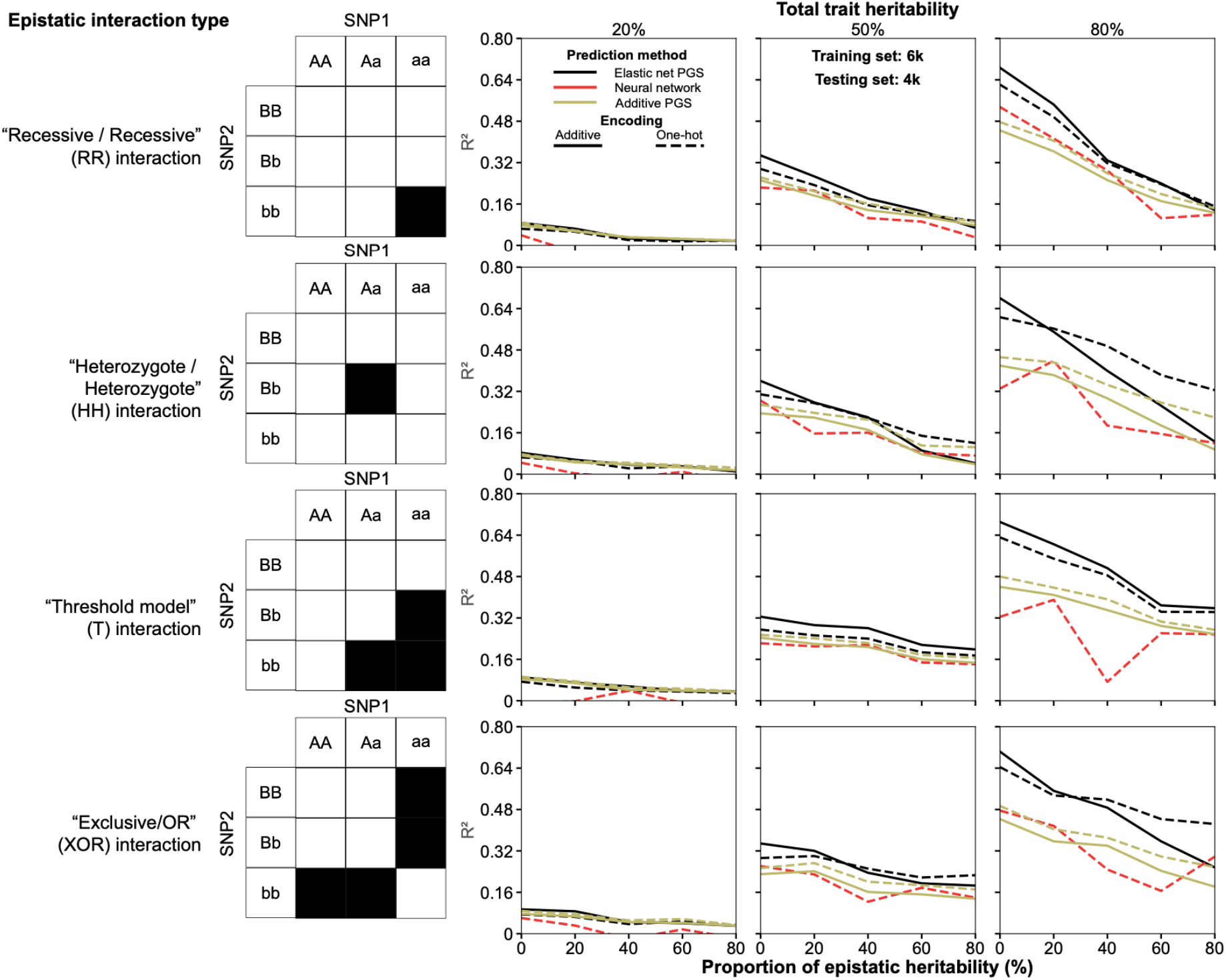
Predictive performances (R^2^) of all PGS methods with a training size of 6k and testing set of 4k samples. Elastic net PGS method is in black, neural network MLP in red and additive P+T PGS in yellow. Solid lines and dashed lines represent additively and one hot encoded models respectively. The plots detail each PGS Method’s performance for a given phenotype. Each row has a different underlying interaction model labelled by the tables; each column has the same total heritability as noted by the column title and within each plot. The *x* axis details the proportion of epistatic contribution for a given trait.

**Supplementary figure 2.**
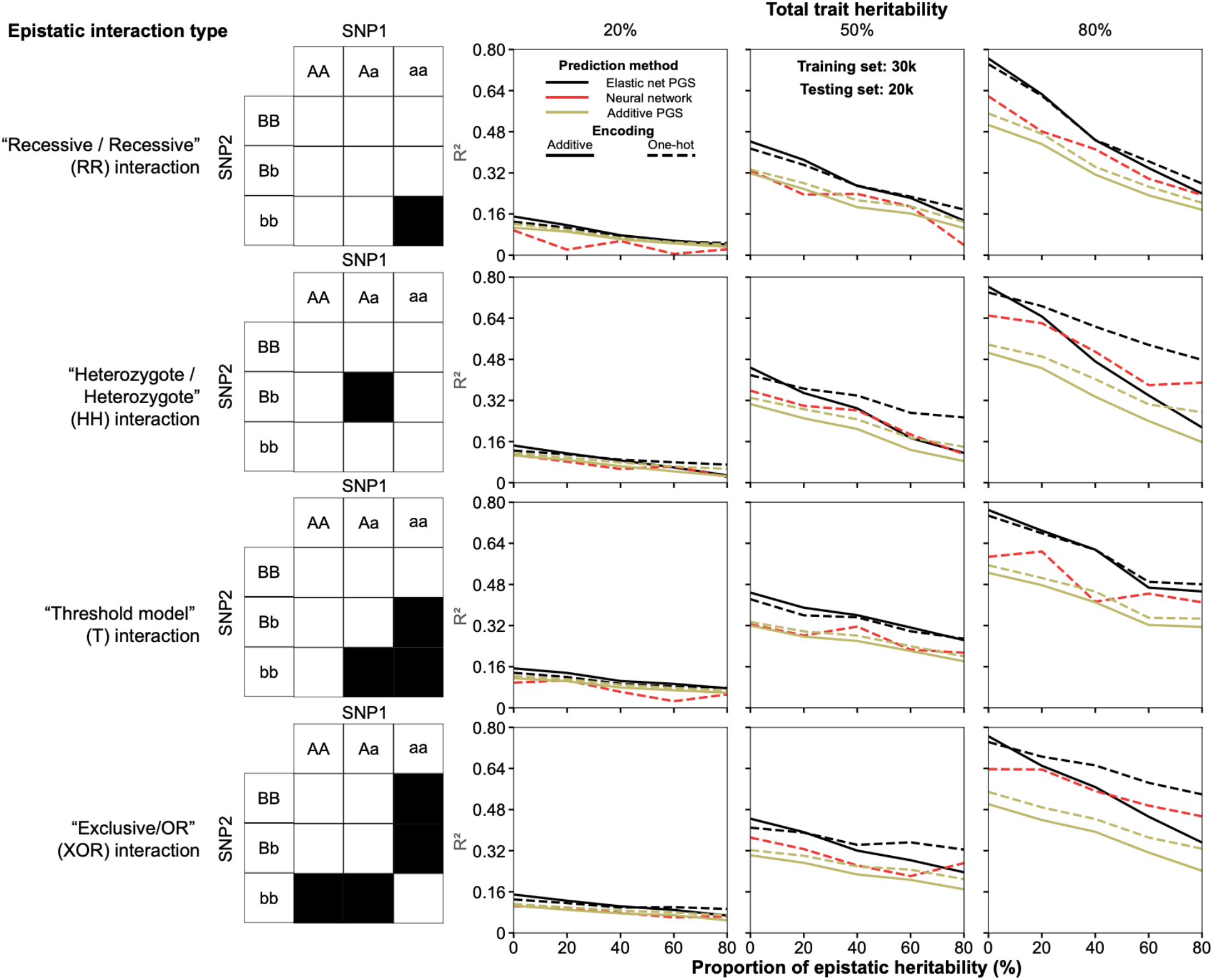
Predictive performances (R^2^) of all PGS methods with a training size of 30k and testing set of 20k samples.

**Supplementary figure 3.**
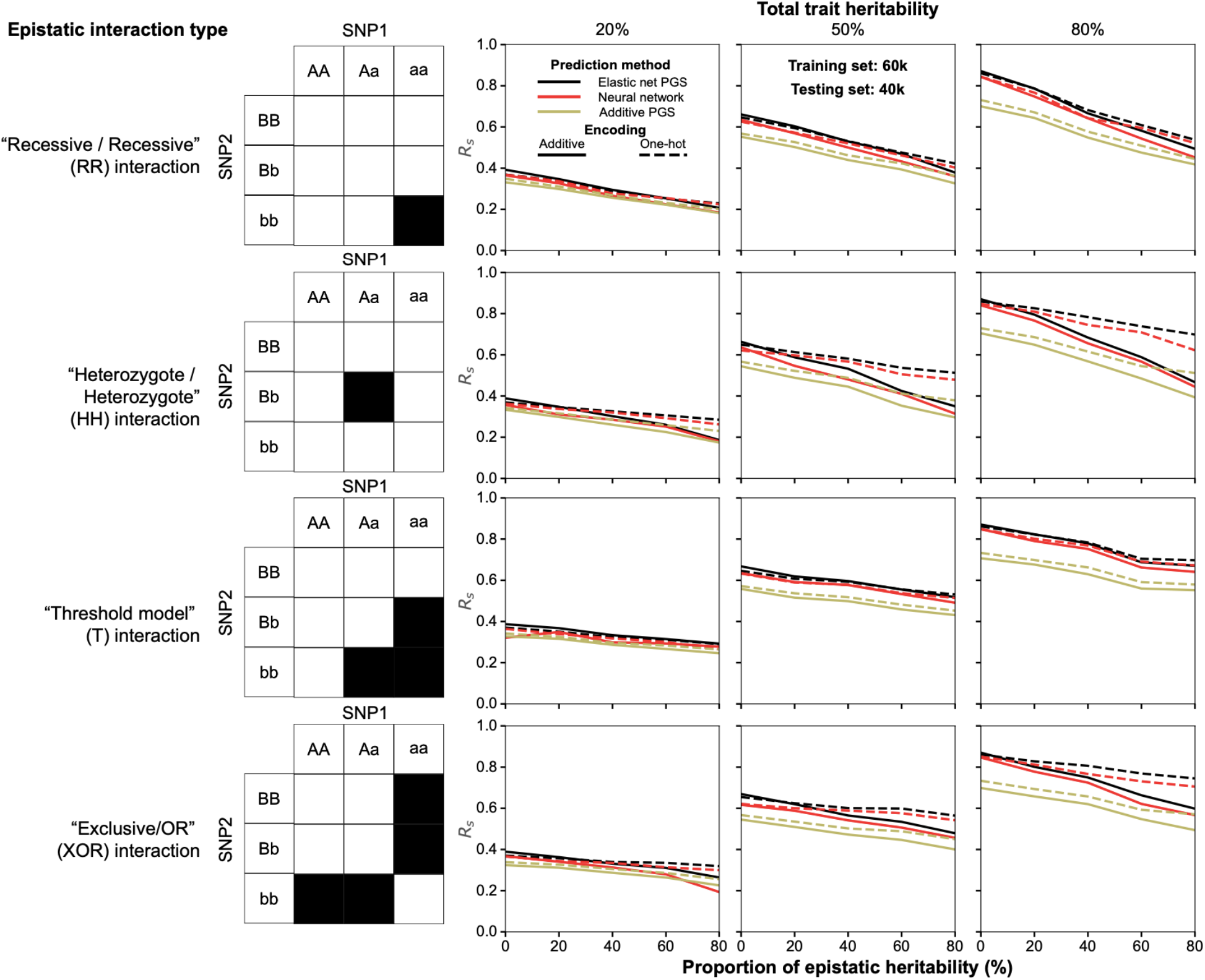
Predictive performances (Spearman R_s_) of all PGS methods with a training size of 60k and testing set of 40k samples.

**Supplementary figure 4.**
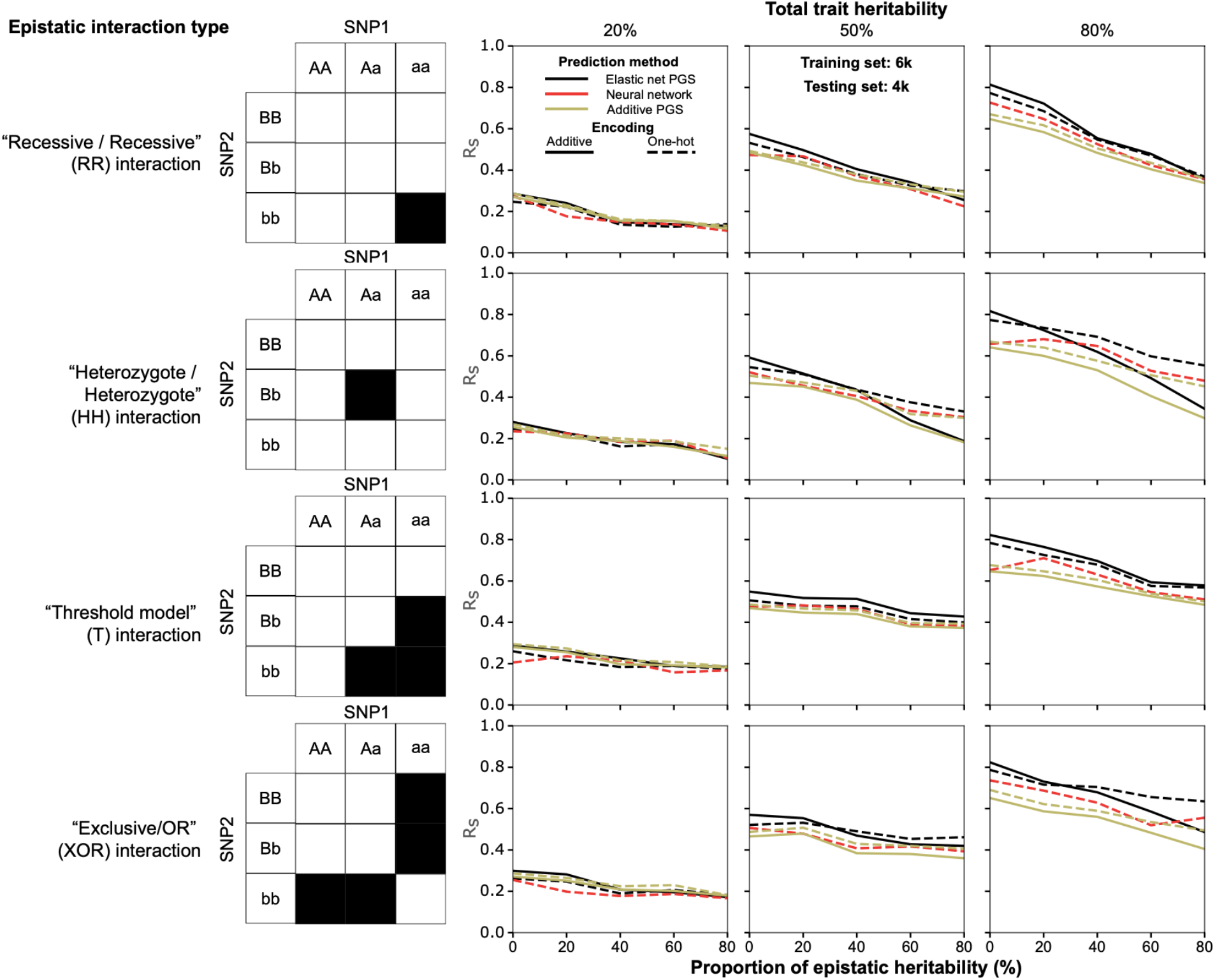
Predictive performances (Spearman R_s_) of all PGS methods with a training size of 6k and testing set of 4k samples.

**Supplementary figure 5.**
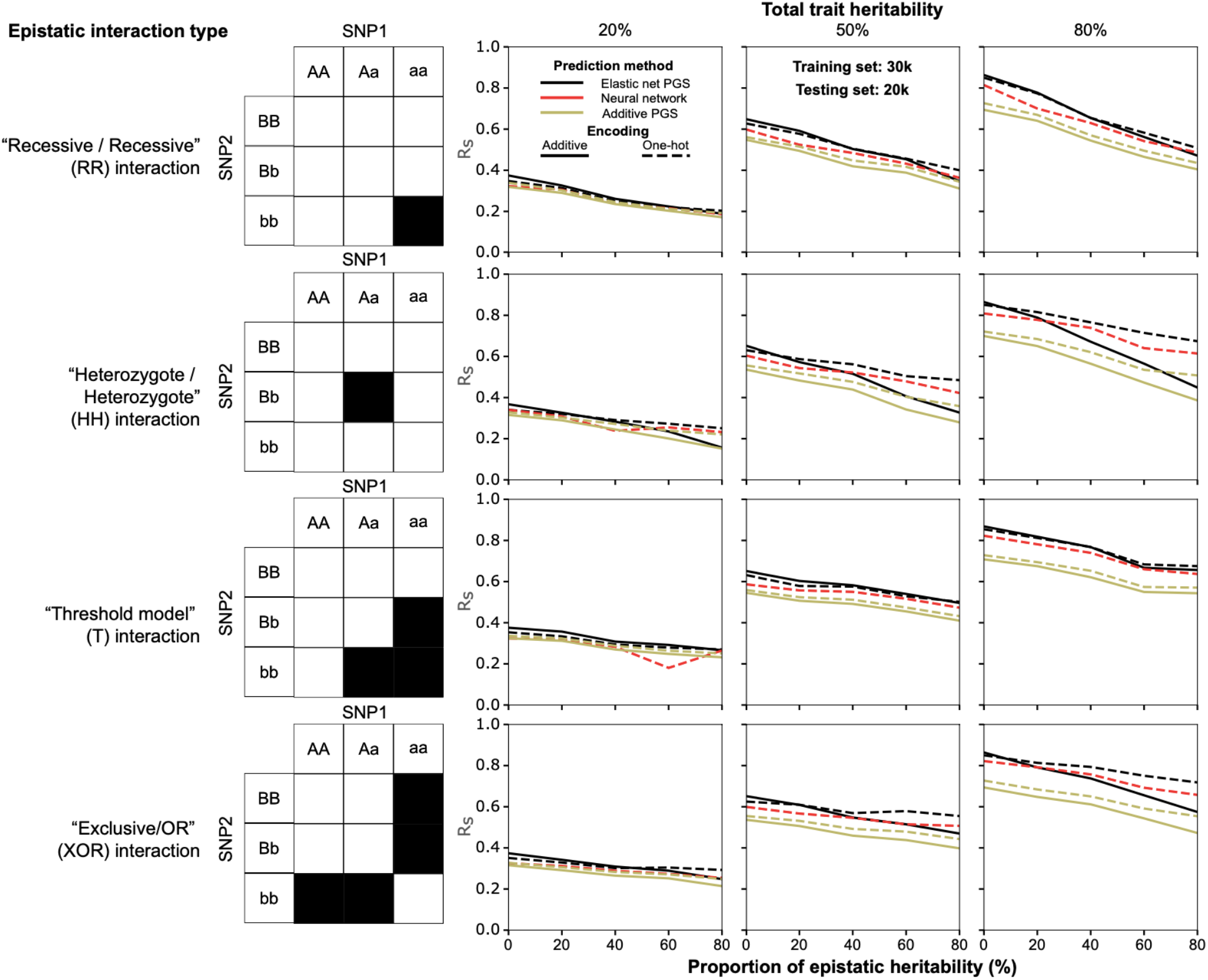
Predictive performances (Spearman R_s_) of all PGS methods with a training size of 30k and testing set of 20k samples.

**Supplementary figure 5.**
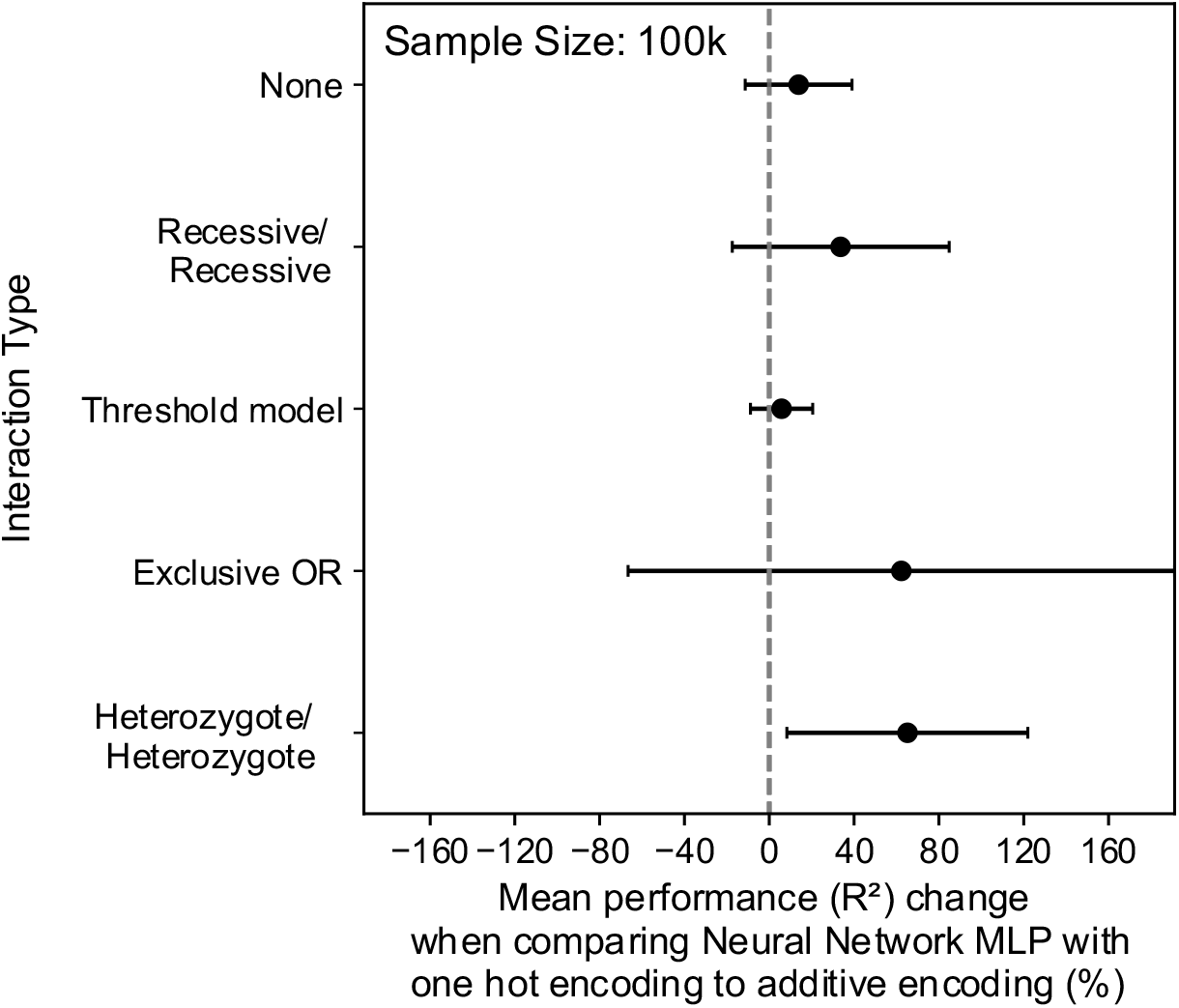
Neural network MLP predicts more accurately with one hot encoded variants. The plot details the percentage change in the predictive performance (R^2^) of neural network MLP method with one hot encoding over that of using additive encoding at the total sample size of 100k, which are measured using the mean and standard deviation of 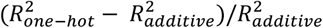 (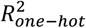 and 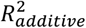 are R^2^ performance of MLP using one hot encoding and additive encoding respectively for a trait) in each trait group by GxG interaction type or with no interactions.

**Supplementary figure 7.**
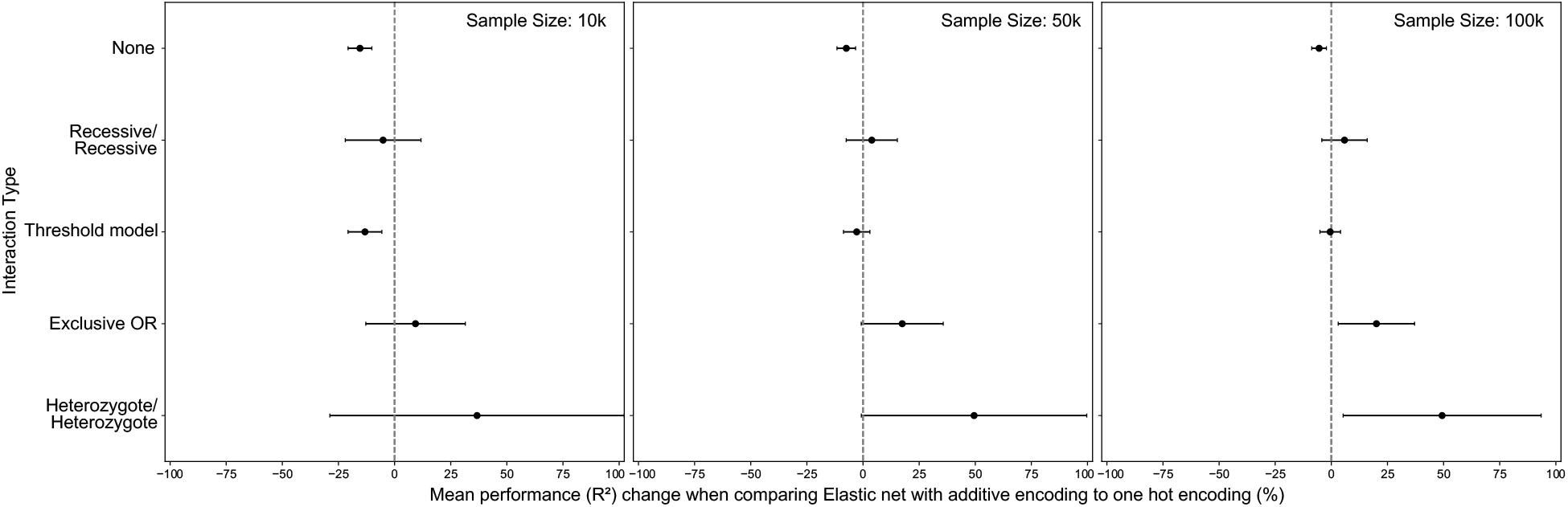
One hot encoding allows elastic net to better predict non-linear traits. The plot details the percentage change in the predictive performance (R^2^) when comparing additively encoded elastic net to one hot encoded elastic net by groups of traits with different interaction types in each of the sample sizes used in this study.

**Supplementary figure 8.**
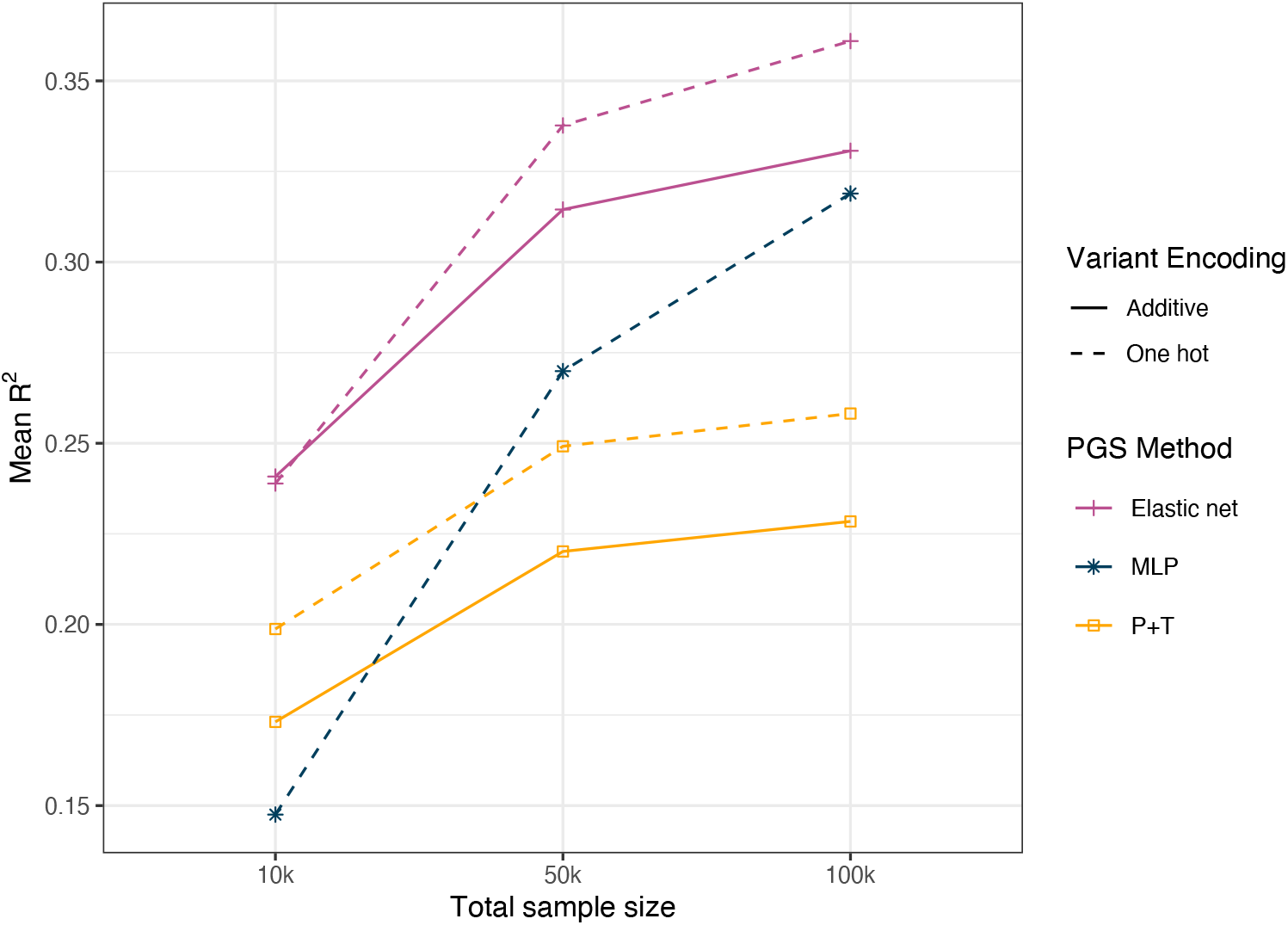
The R^2^ performance improvements of PGS methods by sample size increase. This plot shows the mean R^2^ performance of each PGS method (P+T, elastic net and MLP) with either of the two variant encoding types across all the 60 simulated traits at given sample sizes (10k, 50k, 100k).

**Supplementary figure 9.**
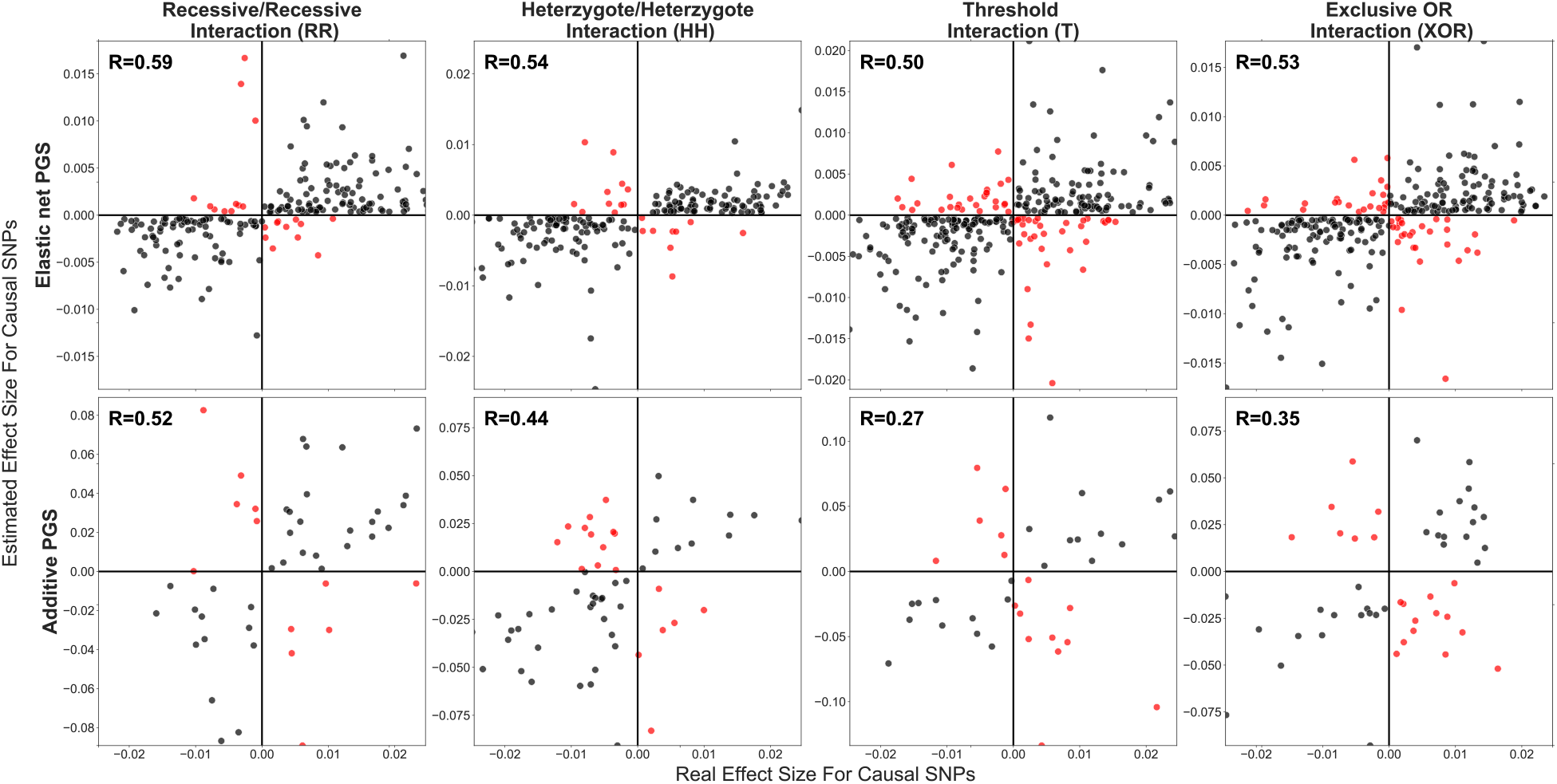
Elastic-net better estimates effect sizes in highly non-additive traits than additive P+T method (80% total heritability and 80% GxG). Each sub-plot corresponds to a single trait under control of different types of interactions, in which the trait has the total heritability of 80% and 80% of heritability is explained by GxG. The total sample size used is 100k. The *x*-axis presents the real linear effects of causal variants and the estimated effect sizes are displayed on the y axis. The upper row shows non-zero effect sizes estimated by Elastic net for causal variants; bottom row shows effect sizes of causal variants estimated by P+T. Columns are separated by the GxG interaction type present in the trait (i.e. XOR, RR, HH and T). Points in the off diagonal are coloured in red. The Spearman correlation between the two effect sizes across all the variants in each plot is labelled at the top left. Note that for clarity any effect size estimated to be exactly zero is removed from the plot.

**Supplementary figure 10.**
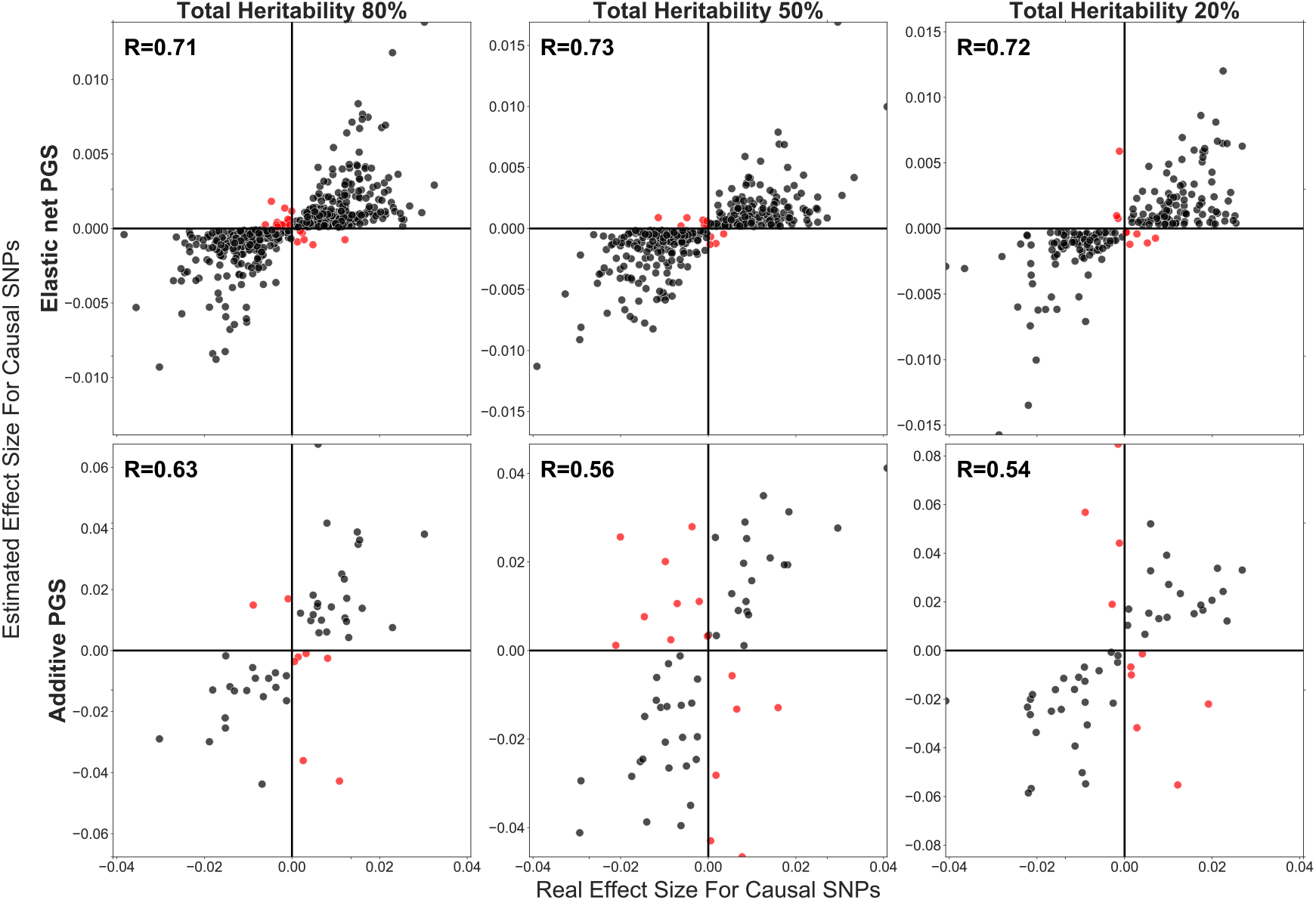
Elastic-net better learns effect sizes of variants in linear traits than additive P+T. Each sub-plot corresponds to a single trait with no interactions (i.e. purely additive traits). The total sample size used is 100k. The x-axis presents the real linear effects for each causal variant (from estimation) and the y axis shows the estimated effect sizes using EN or P+T. The upper row shows non-zero effect sizes of causal variants estimated by Elastic net for traits with 80%, 50%, and 20% total heritability from left to right respectively; bottom row shows effect sizes of causal variants estimated by P+T. Points in the off diagonal are coloured in red. The Spearman correlation between the two effect sizes across all the variants in each plot is labelled at the top left. Note that for clarity any effect sizes estimated to be exactly zero are removed from the plot.

**Supplementary figure 11.**
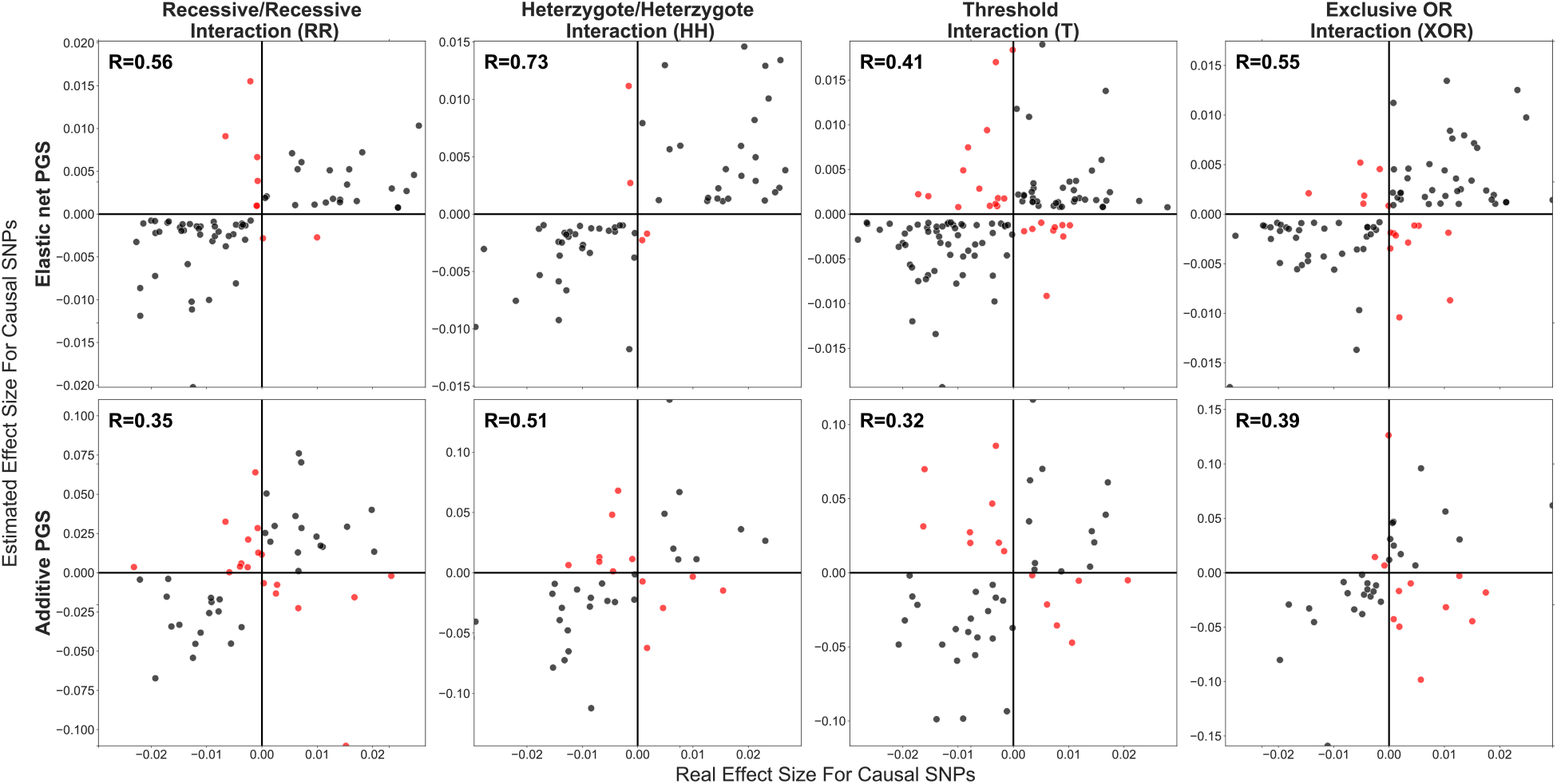
Elastic-net better estimates effect sizes in highly non-additive traits than additive P+T (80% total heritability and 20% GxG). Each sub-plot corresponds to a single trait under control of different types of interactions, in which the trait has the total heritability of 80% and 20% of heritability is explained by GxG. The total sample size used is 100k. The *x*-axis presents the real linear effects of causal variants and the estimated effect sizes are displayed on the *y*-axis. The upper row shows non-zero effect sizes estimated by Elastic net for causal variants; bottom row shows effect sizes of causal variants estimated by P+T. Columns are separated by the interaction type present in the trait (i.e. XOR, RR, HH and T). Points in the off diagonal are coloured in red. The Spearman correlation between the two effect sizes across all the variants in each plot is labelled at the top left. Note that for clarity any effect size estimated to be exactly zero is removed from the plot.

**Supplementary figure 12.**
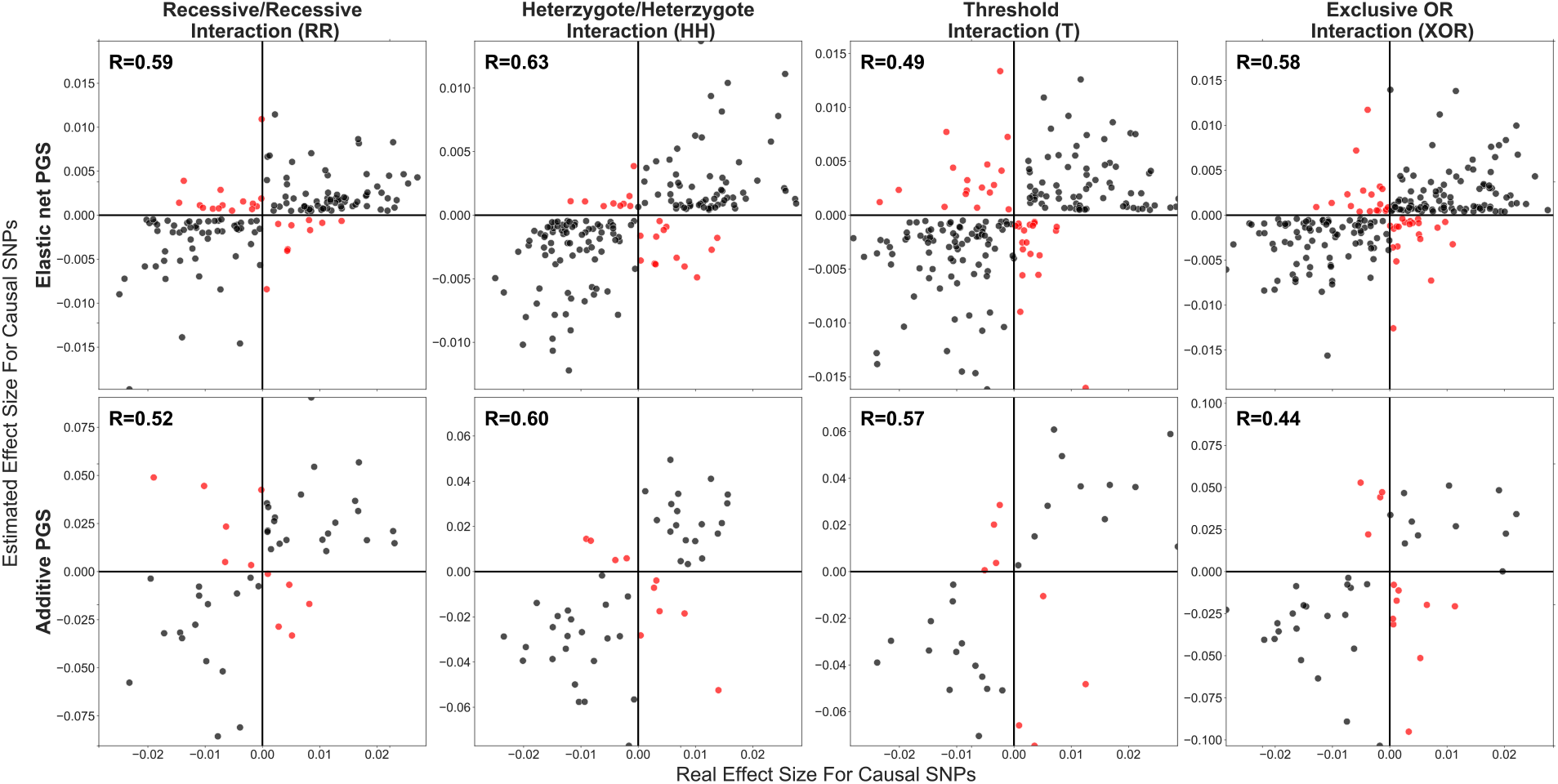
Elastic-net better estimates effect sizes in highly non-additive traits than additive P+T (80% total heritability and 50% GxG interaction). Each sub-plot corresponds to a single trait under control of different types of interactions, in which the trait has the total heritability of 80% and 50% of heritability is explained by GxG. The total sample size used is 100k. The x-axis presents the real linear effects of causal variants and the estimated effect sizes are displayed on the y-axis. The upper row shows non-zero effect sizes estimated by Elastic net for causal variants; bottom row shows effect sizes of causal variants estimated by P+T. Columns are separated by the interaction type present in the trait (i.e. XOR, RR, HH and T). Points in the off diagonal are coloured in red. The Spearman correlation between the two effect sizes across all the variants in each plot is labelled at the top left. Note that for clarity any effect size estimated to be exactly zero is removed from the plot.

**Supplementary figure 13.**
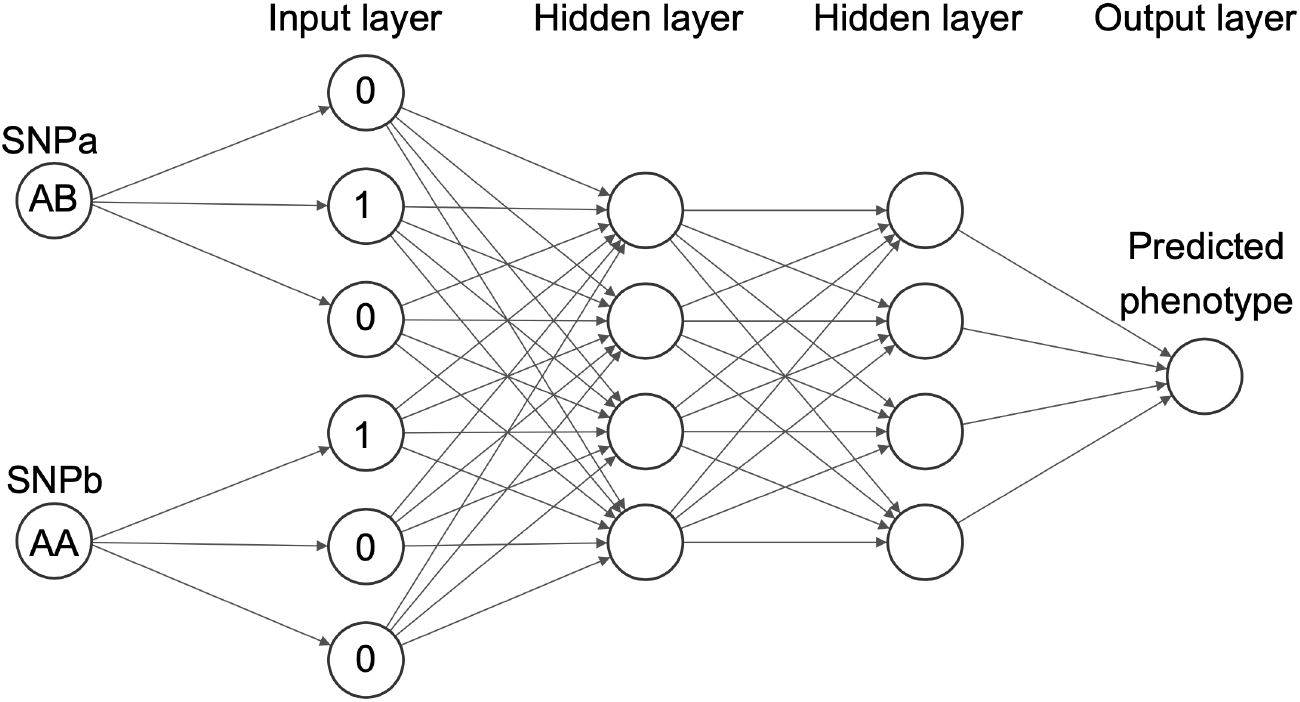
One hot encoded Neural network (NN) schematic. The above is an example of the MLP network structure used in this study. This NN has two input variants, two hidden layers and an output layer where the phenotype is predicted. The input variants are firstly encoded into their genotype classes through one hot encoding, then these are passed through the network’s hidden layers and finally the phenotype is predicted. The NNs in this study had 100,455 variants as input.

**Supplementary figure 14.**
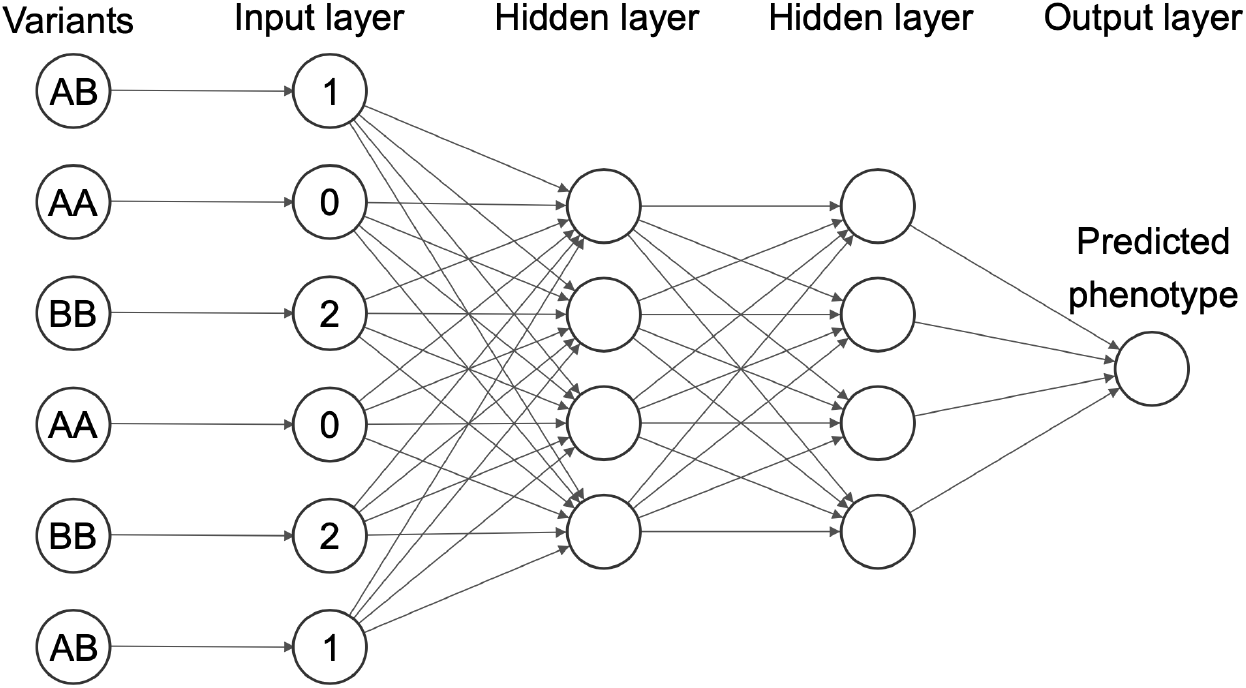
Additively encoded Neural network (NN) schematic. The plot is an example of the MLP network structure used in this study. This NN has six input variants, two hidden layers and an output layer where the phenotype is predicted. The input variants are encoded by counting the number of affect alleles, then these are passed through the network’s hidden layers and finally the phenotype is predicted. The NNs in this study had 100,455 variants as input.

